# Risk of death among teachers in England and Wales during the Covid19 pandemic

**DOI:** 10.1101/2021.02.23.21252143

**Authors:** Sarah J Lewis, Kyle Dack, Caroline L Relton, Marcus R Munafò, George Davey Smith

## Abstract

**Objectives:** To estimate occupation risk from Covid19 among teachers and others working in schools using publicly available data on mortality in England and Wales.

**Design:** Analysis of national death registration data from the Office for National Statistics.

**Setting:** England and Wales, March 20^th^ to 28^th^ December 2020, during the Covid19 pandemic.

**Participants:** The total working age population in England and Wales plus those still working aged over 65.

**Primary and Secondary outcomes:** Death with Covid19 as a primary outcome and death from all causes as a secondary outcome.

**Results:** Across occupational groups there was a strong correlation between Covid19 mortality and both non-Covid19 and all-cause mortality. The absolute mortality rates for deaths with Covid19 were low amongst those working in schools (from 10 per 100,000 in female primary school teachers to 39 per 100,000 male secondary school teachers) relative to many other occupations (range: 10 to 143 per 100,000 in men; 9 to 50 per 100,000 in women).There was weak evidence that secondary school teachers had slightly higher risks of dying with Covid19 compared to the average for all working aged people, but stronger evidence of a higher risk compared to the average for all professionals; primary school teachers had a lower risk. All-cause mortality was also higher amongst all teachers compared to all professionals. Teaching and lunchtime assistants were not at higher risk of death from Covid19 compared with all working aged people.

**Conclusion:** There was weak evidence that Covid19 mortality risk for secondary school teachers was above expectation, but in general school staff had Covid19 mortality risks which were proportionate to their non-covid mortality risk.

**Strengths and limitations of this study:**

- We used routinely collected data on all deaths in England and Wales;, which are near-complete and not susceptible to serious ascertainment biases.
- We were able to compare mortality data for teachers and other school workers with all other occupational groups and with the working-age population.
- The number of deaths due to Covid19 were small and thus differences between the specific occupational groups were imprecisely estimated.
- We did not have access to individual level mortality data so were not able to account for potential confounders such as comorbidities or household size.
- For those working in school aged over 65 we had neither mortality rates per 100,000 nor total numbers within the group; we only had number of deaths and a 5-year average and we do not know whether the denominators have changed for this group over the last 5 years.

## Background

School closures have been implemented in many countries in an effort to slow the spread of the SARS-CoV-2 virus responsible for the Covid19 pandemic. This has affected hundreds of millions of children globally and has been contentious due to harms that have arisen to children as a result of school closures (1). Schools in England and Wales closed to most pupils (except a small number of children of critical workers and some vulnerable children) on the 20^th^ March 2020 and did not reopen fully until September 2020; they closed again for the Christmas holidays (between 11^th^ and 18^th^ December) and reopened on the 8^th^ March in England, although at the time of writing (15^th^ March) secondary schools in Wales are still closed.

Whether or not teachers and others working in schools are at higher risk from Covid19 as a result of schools being open is central to decisions on school reopening, but until recently there has not been good data on this. Two as yet unpublished record linkage studies carried out in Scotland (2) and Finland (3) show that teachers have a greater risk of being diagnosed as a Covid19 case compared with the general population. There was a 25% higher risk in the Scottish study, and between 50% and 70% higher risk in the Finnish study. A study in Sweden found that teachers delivering in person teaching had a two-fold higher risk of Covid-19 infection compared to those who were teaching remotely (4). However, studies examining transmission in schools have found this to occur at low levels (5,6). One reason for the discrepancy is that teachers are likely to be under greater surveillance, and therefore undergo more testing than those working in other occupations. In the Scottish study teachers were much more likely to be tested over the autumn term compared with the general population, and this is likely to explain at least some the elevated number of cases among teachers.

Analyses of more robust outcomes such as hospitalisation from Covid19 are less susceptible to detection bias and may be more reliable. The Scottish teachers study (2) found that 47 out of 11,101 teachers were hospitalised with Covid19 up to 26^th^ November 2020, with an odds ratio (OR) of 0.98 (95% CI 0.67 to 1.45) for hospitalisation compared with controls matched on age, sex and general practice registration and adjusting for ethnicity, deprivation, comorbidity counts and sharing a household with a healthcare worker. Sweden has been an outlier in the Covid19 pandemic in that schools have remained open for children up to 16 years of age. A study by the Swedish Public Health Agency found that 20 out of 103,596 schoolteachers in Sweden received intensive care treatment up to the 30^th^ June 2020, an age and sex matched analysis comparing teachers with other occupations (except health care workers) reported an OR of 0.43 (95% CI 0.28 to 0.68) (7). Similarly, a study carried out in Norway did not find higher rates of hospitalisation with Covid19 for school teachers, after adjustment for age, sex and country of birth (8). There is an absence of evidence on the risk to teaching and lunchtime assistants working in schools.

The Office for National Statistics (ONS) routinely collect mortality data for those living in England and Wales. They have recently released data on deaths with Covid19 and all-cause mortality by occupation (9). We used this data to compared mortality risk among school workers and all working aged adults in England and Wales. We also compared mortality rates among teachers to all professionals, because there are large differences in mortality risk by occupation, largely driven by differences in socioeconomic circumstances (10).

## Methods

The ONS mortality statistics used for this analysis are based on mandatory registration of all deaths in England and Wales. So far 3 datasets on mortality with Covid19 by occupation have been released; the first covered the period from 8^th^ March to the 20^th^ April which included the peak of wave 1 deaths, the second covered the period between 8^th^ March to the 25^th^ May and the final dataset (the one we used for the analysis in this paper) covered the period from 8^th^ March to the 28^th^ December.

We used these data to determine whether the risk of death from Covid19 was higher among teachers and others working in the education sector than those working in other occupations. We compared the number of deaths with historical data on deaths for the same occupational group over 5 years (2015 to 2019) and compared death rates among teachers, teaching and lunchtime assistants death rates among all working aged individuals over the same period of the pandemic. We also compared mortality among teachers with all professionals. Finally, we investigated the ratio of mortality with Covid19 to mortality from other causes among different occupational groups to determine whether school staff were outliers.

### Occupational exposure groups

Occupation was reported on the death certificate at the time of death registration by the informant and was coded according to the ONS standard occupational classification 2010 (10). Population counts for occupational groups were estimated from the Annual Population Survey (APS), conducted in 2019 (11). The APS is the largest ongoing household survey in the UK, conducting interviews with members of randomly selected households.

The group ‘teaching and educational professionals’, as defined by the ONS, includes; higher education teaching professionals, further education teaching professionals, secondary education teaching professionals, primary and nursery education teaching professionals, special needs education teaching professionals, senior professionals of educational establishments, education advisers and school inspectors and teaching and other educational professionals not elsewhere defined.

The group ‘teaching and educational professionals’ does not include: teaching assistants, school secretaries, lunchtime assistants or educational support assistants, but these occupations were considered along with teachers working in schools where sufficient data were available.

The group defined by ONS as midday and crossing patrol occupations, includes some individuals who work as road crossing patrols, however not all schools have these and if they do usually only have one, so the group will mainly comprise of those who supervise children in school during their lunch break and who prepare school lunch. Therefore midday and crossing patrol occupations will be shorted and referred to in this manuscript as lunchtime assistants.

### Outcome definition

ONS define deaths with Covid19 as those where Covid19 or suspected Covid19 was mentioned anywhere on the death certificate, including in combination with other health conditions. If a death certificate mentions Covid19, it will not always be the main cause of death but may be a contributory factor.

### Statistical analysis

We calculated the relative risk of death for education professions compared to all working aged (20 to 64 year olds) people and compared teachers to all professionals in England and Wales stratified by sex, for this we used published ONS data on total deaths and age adjusted mortality rates per 100,000 for each group. We derived the denominator (total population) for each group of interest as

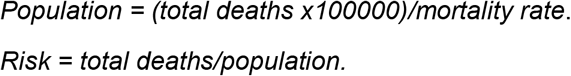

We calculated 95% confidence intervals and *p-*values using the formula;

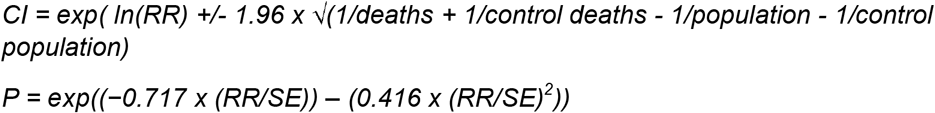

The ONS data included deaths for 8 subgroups of educational professionals, we included those with a sufficient number of Covid19 deaths (≥10) in our analysis. In addition, sufficient data were available to include female teaching and lunchtime assistants in this analysis. We also performed a fixed effect meta-analysis across the 4 occupational groups working in schools among females.

We estimated mortality rates for all other causes (other than Covid19) by calculating the difference between all-cause mortality and mortality with Covid19 for each minor occupational group as defined by ONS (12).

We also produced scatterplots of ‘all cause’ versus ‘with Covid19’ mortality and ‘all other causes’ versus ‘with Covid19*’* mortality by occupational group for men and women separately and calculated Pearson’s correlation coefficient for these plots.

We also calculated proportionate mortality ratios as mortality rates from Covid19 divided by all-cause mortality for occupations working in schools.

Finally we used our estimates from above for the SE and the denominators for each occupational group to estimate the variance for Covid19 mortality. We then performed a weighted least squares regression analysis (weighted by 1/variance) of Covid19 mortality against other cause to determine the increase in Covid19 for 1 death increase in other cause mortality across occupational groups.

## Results

### Deaths from Covid19 and all causes among education professionals and others working in schools

Between the 9^th^ March 2020 and 28^th^ December 2020 there were 68,757 deaths from all causes among the working age (20-64 years) population in England and Wales, 7,961 (12%) of these deaths involved Covid19, of which, 2,494 had occurred prior to the 20^th^ April, 2,267 occurred between 20^th^ April and the 25^th^ May, and 3,200 occurred between the 25^th^ May and the 28^th^ December 2020.

There were 1,326 deaths from all causes among educational professionals during this period; 139 of those deaths were thought to involve Covid19.

**Table 1** shows that during the 10 months covered by the ONS data all-cause and Covid19 mortality rates among educational professionals were lower than for all working aged adults and similar to those for all professionals.

**Table 1.** Age adjusted all cause and with Covid19 mortality per 100,000 population (95% confidence intervals.

| Occupational group | Cause of Mortality | Males |  |  | Females |  |  |
| --- | --- | --- | --- | --- | --- | --- | --- |
|  |  | 8<sup>th</sup> March-20<sup>th</sup> April | 8<sup>th</sup> March-25<sup>th</sup> May | 8<sup>th</sup> March-28<sup>th</sup> December | 8<sup>th</sup> March-20<sup>th</sup> April | 8<sup>th</sup> March-25<sup>th</sup> May | 8<sup>th</sup> March-28<sup>th</sup> December |
| Working aged adults | With Covid19 | 9.9<br>(9.4-10.4) | 19.1<br>(18.4-19.8) | 31.4<br>(30.6-32.3) | 5.2<br>(4.9-5.6) | 9.7<br>(9.3-10.2) | 16.8<br>(16.2-17.5) |
|  | All cause | 41.6<br>(40.6-42.6) | 78.1<br>(76.7-79.4) | 256.0<br>(253.5-258.4) | 26.3<br>(25.5-27.1) | 48.4<br>(47.3-49.4) | 158.3<br>(156.4-160.2) |
|  | Ratio* | 1:4.2 | 1:4.1 | 1:8.2 | 1:5.1 | 1:5.0 | 1:9.4 |
| Professional Occupations | With Covid19 | 5.6<br>(4.6-6.6) | 11.6<br>(10.2-13.0) | 17.6<br>(15.9 - 19.3) | 4.2<br>(3.3-5.2) | 8.0<br>(6.8 to 9.3) | 12.8<br>(11.2- 14.4) |
|  | All cause | 22.2<br>(20.2-24.1) | 41.2<br>(38.5-43.8) | 130.4<br>(125.7-135.2) | 22.0<br>(19.9-24.1) | 39.3<br>(36.5-42.1) | 120.0<br>(115.2-124.9) |
|  | Ratio* | 1:4 | 1:3.6 | 1:7.4 | 1:5.2 | 1:4.9 | 1:9.4 |
| Educational professionals | With Covid19 | 6.7<br>(4.1-10.3) | 12.9<br>(9.3-17.4) | 18.4<br>(14.0-23.6) | 3.3<br>(2.0-4.9) | 6.0<br>(4.2-8.1) | 9.8<br>(7.5-12.5) |
|  | All cause | 28.0<br>(22.4-34.5) | 48.1<br>(40.4-55.7) | 153.4<br>(139.8-167.1) | 20.4<br>(16.9-24.0) | 37.2<br>(32.5-42.0) | 110.0<br>(101.9-118.2) |
|  | Ratio* | 1:4.2 | 1:3.7 | 1:8.3 | 1:6.2 | 1:6.2 | 1:11.2 |
| *Ratio of deaths involving Covid19 deaths to all deaths |  |  |  |  |  |  |  |

Covid19 was involved in 1 in 8 male deaths and 1 in 9 female deaths among working aged people; among education professionals these figures were approximately 1 in 8 deaths among males and 1 in 11 among females. Covid19 was involved in a higher proportion of deaths early on in the pandemic; data covering the 6 weeks of the first peak in the UK showed that Covid19 was involved in 1 in 4 deaths in males and 1 in 6 deaths in female education professionals during this time.

We examined number of deaths by occupation among those working in schools aged 20-64 years (Supplementary table S1) and among those aged 65 or older (Supplementary table S2). Among working aged people when deaths across all occupations working in schools were combined, we observed 15% more deaths (excess) compared with the 2015-2019 average for the same period among men and 5% more among women. For women this was less than the excess observed across all occupations and among all professionals; for men it was similar. Deaths with Covid19 appeared to account for almost all the excess among men whereas among women there were twice as many deaths with Covid19 among school workers as there were excess deaths.

Among the older age group there were much higher numbers of deaths compared with their 5-year average (74% more deaths amongst males and 37% among females). However, only 33% of excess deaths in men and 37% in women were thought to involve Covid19. The group of all professionals and of all occupations had fewer excess deaths as a proportion of their 5-year average but a higher proportion of the deaths involved Covid19 compared with those working in schools.

### Comparison of Covid19 and all-cause deaths among education professionals and those working in schools compared to the working aged population

**Table 2** shows that compared to the total working aged population there was strong evidence that the risk of dying with Covid19 was lower among all education professionals combined (RR_Males_ 0.59, 95%CI 0.46 to 0.75, RR_females_ 0.58, 95%CI 0.46 to 0.73). There was also some evidence of a lower risk of death with Covid19 among female primary and nursery education professionals (RR 0.60, 95%CI 0.38 to 0.93). For both male and female secondary school teachers the relative risk estimate shows a 25% higher risk of death from Covid19, although there was a great deal of uncertainty around these estimates (RR_Males_ 1.25, 95%CI 0.87 to 1.80, RR_females_ 1.26, 95%CI 0.84 to 1.90). There was little evidence of a difference in risk between teaching assistants, or lunchtime assistants compared with working aged people. A fixed effect meta-analysis across the 4 occupational groups working in schools in women showed that risk was similar to all working aged people (RR 0.95, 95CI 0.77 to 1.14), with weak evidence of heterogeneity across the groups (I^2^=55.2%, Chi^2^=6.697, p=0.08).

**Table 2.** **Covid19 deaths for educational professions aged 20-64 years compared to all working aged people, between 9th March and 28th December 2020**

| Description | Men |  |  |  |  | Women |  |  |  |  |
| --- | --- | --- | --- | --- | --- | --- | --- | --- | --- | --- |
|  | Risk* | Risk for all workers | RR | 95% CI | P-value | Risk* | Risk for all workers | RR | 95% CI | P-value |
| All educational professionals | 0.00018 | 0.00031 | 0.59 | 0.46 - 0.75 | 0.000018 | 0.000098 | 0.00017 | 0.58 | 0.46 - 0.73 | 0.0000058 |
| Higher education teaching professionals | 0.00012 | 0.00031 | 0.37 | 0.20 - 0.68 | 0.0016 | NA<sup>1</sup> |  |  |  |  |
| Further education teaching professionals | 0.00025 | 0.00031 | 0.79 | 0.42 - 1.46 | 0.46 | NA<sup>1</sup> |  |  |  |  |
| Secondary education teaching professionals | 0.00039 | 0.00031 | 1.25 | 0.87 - 1.80 | 0.23 | 0.00021 | 0.00017 | 1.26 | 0.84 - 1.90 | 0.27 |
| Primary and nursery teaching professionals | NA<sup>1</sup> |  |  |  |  | 0.00010 | 0.00017 | 0.60 | 0.38 - 0.93 | 0.024 |
| Teaching assistants | NA<sup>1</sup> |  |  |  |  | 0.00015 | 0.00017 | 0.89 | 0.65 - 1.23 | 0.5 |
| Lunchtime assistants | NA<sup>1</sup> |  |  |  |  | 0.00019 | 0.00017 | 1.14 | 0.72 - 1.82 | 0.58 |
| *Total deaths divided by denominator <sup>1</sup>Less than 10 deaths, rate not reported. |  |  |  |  |  |  |  |  |  |  |

**Table 3** shows strong evidence that the risk of death from all causes during this pandemic period was lower for higher and further education teacher professionals compared with the working aged population of England and Wales. The was also evidence that male secondary school teachers (RR 1.26, 95%CI 1.11 to 1.43) and female primary school teachers (RR 1.18, 95%CI 1.06 to 1.32) had slightly higher mortality risk compared to the general population.

**Table 3.**
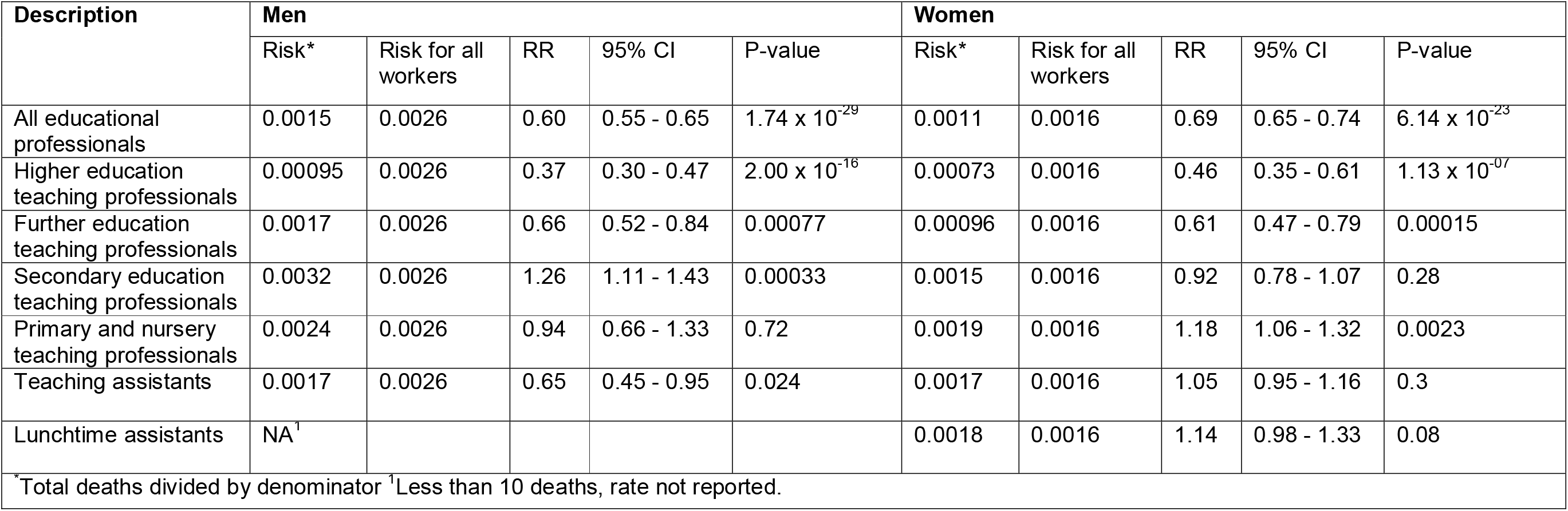
**All-cause mortality for school staff aged 20-64 years compared to all working aged people, between 9th March and 28^th^ December 2020**

| Description | Men |  |  |  |  | Women |  |  |  |  |
| --- | --- | --- | --- | --- | --- | --- | --- | --- | --- | --- |
|  | Risk* | Risk for all workers | RR | 95% CI | P-value | Risk* | Risk for all workers | RR | 95% CI | P-value |
| All educational professionals | 0.0015 | 0.0026 | 0.60 | 0.55 - 0.65 | 1.74 x 10<sup>-29</sup> | 0.0011 | 0.0016 | 0.69 | 0.65 - 0.74 | 6.14 x 10<sup>-23</sup> |
| Higher education teaching professionals | 0.00095 | 0.0026 | 0.37 | 0.30 - 0.47 | 2.00 x 10<sup>-16</sup> | 0.00073 | 0.0016 | 0.46 | 0.35 - 0.61 | 1.13 x 10<sup>-07</sup> |
| Further education teaching professionals | 0.0017 | 0.0026 | 0.66 | 0.52 - 0.84 | 0.00077 | 0.00096 | 0.0016 | 0.61 | 0.47 - 0.79 | 0.00015 |
| Secondary education teaching professionals | 0.0032 | 0.0026 | 1.26 | 1.11 - 1.43 | 0.00033 | 0.0015 | 0.0016 | 0.92 | 0.78 - 1.07 | 0.28 |
| Primary and nursery teaching professionals | 0.0024 | 0.0026 | 0.94 | 0.66 - 1.33 | 0.72 | 0.0019 | 0.0016 | 1.18 | 1.06 - 1.32 | 0.0023 |
| Teaching assistants | 0.0017 | 0.0026 | 0.65 | 0.45 - 0.95 | 0.024 | 0.0017 | 0.0016 | 1.05 | 0.95 - 1.16 | 0.3 |
| Lunchtime assistants | NA<sup>1</sup> |  |  |  |  | 0.0018 | 0.0016 | 1.14 | 0.98 - 1.33 | 0.08 |

\*Total deaths divided by denominator <sup>1</sup>Less than 10 deaths, rate not reported.

We also calculated relative risks for mortality with Covid19 and all-cause mortality in educational professionals versus all professionals (Supplementary Tables S3 and S4). We found strong evidence that both male and female secondary school teachers had a higher risk of mortality with Covid19 compared with all professionals. In addition male and female primary and secondary school teachers had higher mortality risks for all causes compared with all professionals.

### Comparison of Covid19 to other cause mortality across occupational groups

**Figures 1 and 2** show that there are strong correlations between all other cause mortality and mortality with Covid19. Weighted least squares regression analysis showed that for every death from other causes there were 0.11 (95%CI=0.09 to 0.13, *p*=2.0×10^−16^) deaths from Covid19, with no meaningful difference between men (0.11, 95%CI 0.09 to 0.13, *p*=2.49×10^−12^) and women (0.10, 95%CI 0.08 to 0.13, *p*=4.41×10^−9^).

**Figure 1.**
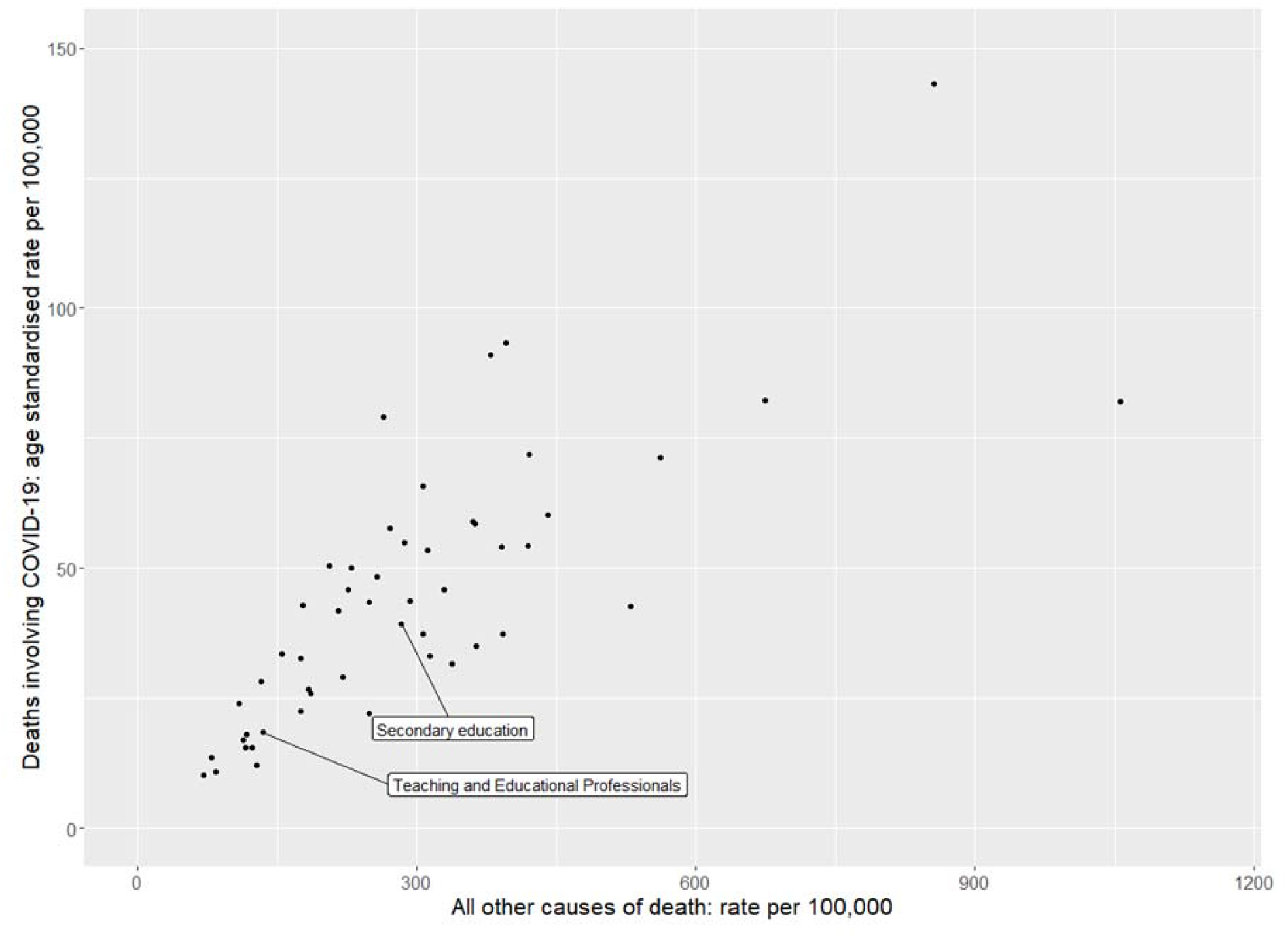
**Scatterplot of mortality rate (per 100,000) for Covid19 and all other causes of death for men, between 9th March and 28th December 2020** Pearson’s correlation coefficient = 0.78, *p* = 1.4 × 10^−11^

**Figure 2.**
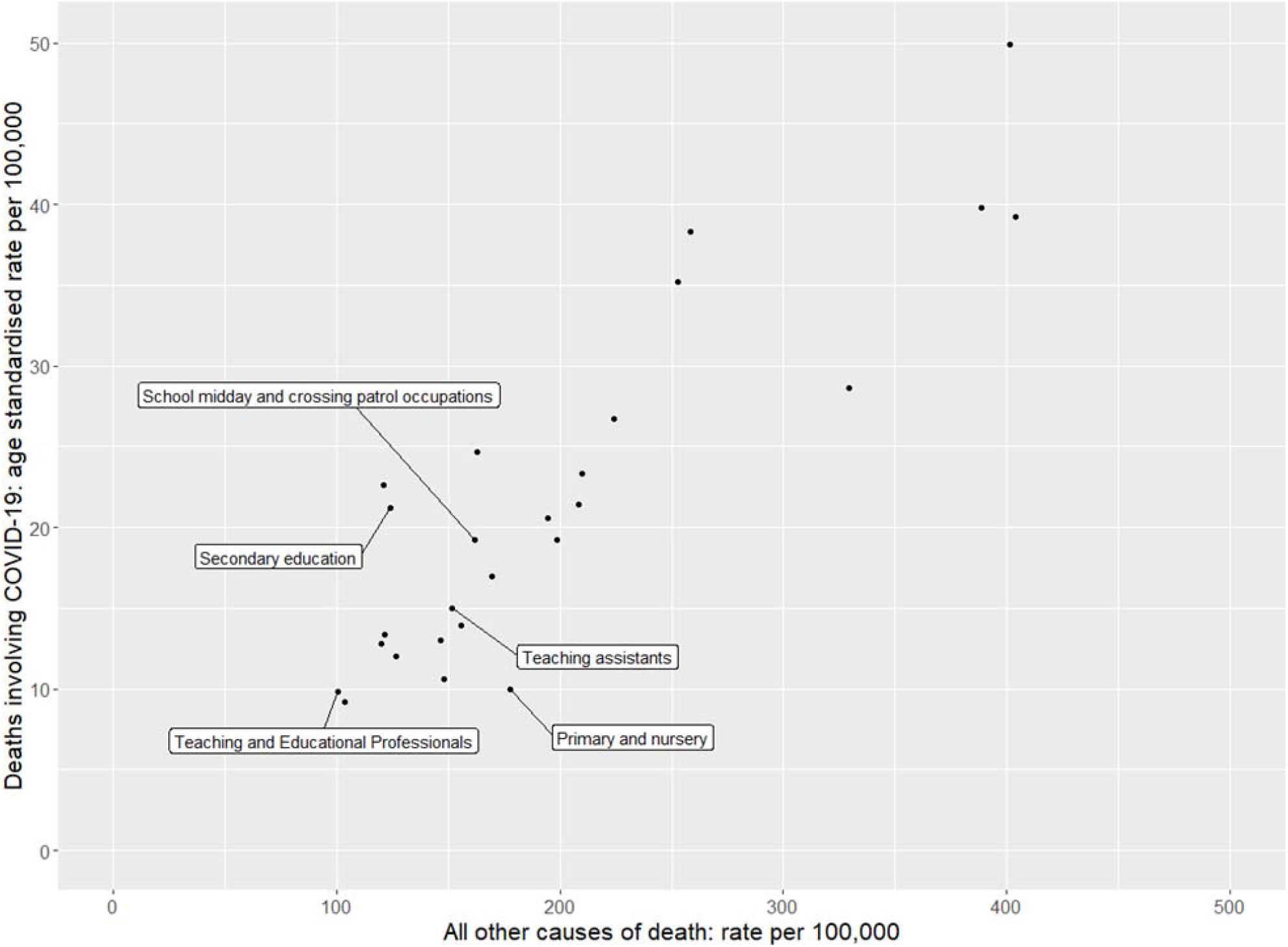
**Scatterplot of mortality rate (per 100,000) for Covid19 and all other causes of death for women, between 9th March and 28th December 2020** Pearson’s correlation coefficient = 0.88, *p* = 1.7. × 10^−09^

We also plotted mortality with Covid19 versus all-cause mortality on separate scatterplots (Figures S1 and S2) and found similar results.

### Ratio of mortality with Covid19 to other causes

The ratio of deaths with Covid19 to other causes of death, ranged from 1:3 among male nurses and midwifes to 1:13 among male elementary construction occupations (Supplementary Table S5). The ratio of Covid19 to other causes of death for male secondary school teachers was 1:7, which was the same as the average for all male professionals and all working aged men. Among women (Supplementary Table S6), the ratio of mortality with Covid19 to other causes of death, ranged from 1:5 among welfare professionals (social workers and probation officers) to 1:18 among primary and nursery schoolteachers; ratios for female secondary school teachers, teaching assistants and lunchtime assistants were 1:6, 1:10 and 1:8 respectively. Proportionate mortality rates (Covid19 divided by all-cause mortality are presented in Supplementary Table S7.

## Discussion

### Covid19 deaths among education professionals and others working in schools

We used routinely collected data on mortality for the Office of National Statistics in the UK to study deaths with Covid19 among teachers and other school staff, during the pandemic in 2020. We found that all educational professionals combined had fewer deaths per 100,000 occurring with Covid19 compared to the general population. This is to be expected since the total working aged population of England and Wales will include those who are unable to work due to ill health. The group ‘education professionals’ includes those working in universities and school inspectors, many of whom have been working from home throughout the pandemic. We therefore investigated excess deaths specifically among school teachers and other staff working in schools and compared them to all those currently working and all professionals. There were fewer deaths than the 5-year average among female primary and secondary school teachers and the number of deaths among male teachers were similar to the 5-year average. There were more deaths among teaching assistants compared to the 5-year average, but only around half of the excess deaths were thought to involve Covid19. For all women working in schools combined there was a small increase in the number of deaths (5%) compared with the 5-year average, but the number of deaths occurring with Covid19 was greater than the excess, suggesting that some of those dying with Covid19 may have died over the course of a normal year from other causes. For those working in schools who were aged 65 and over there were large excesses in deaths compared with the average for the previous 5 years (74% for men and 37% for women), but only around a third of the excess deaths were thought to involve Covid19. It is possible that some excess deaths, were due to undiagnosed Covid19 among those who did not have classic symptoms (13), but this is less likely for later deaths as the amount of testing for Covid19 increased dramatically over time in the UK. We do not have information on other causes of death within this population over this time period, however in 2019 the most common causes of death among over 65s in England and Wales were heart disease and cancer (14). It is possible that more school staff were dying from other causes during this period due to delayed treatments for other conditions or due to an unwillingness to seek help for fear of contracting Covid19 or of overburdening the healthcare system. It is also possible the number of school staff in this group has increased over the last 5 years.

### Relative risk of death with Covid19 among school staff

We found that secondary school teachers had around a 25% higher risk (in both men and women) of dying with Covid19 compared with the general population, although the evidence was weak. There was stronger evidence of an increased risk to secondary school teachers compared to all professionals. However, all-cause mortality was also increased among secondary school teachers compared to all professionals.

Proportionate mortality ratios (mortality from a specific cause: ‘all cause’ or ‘all other cause mortality’) are routinely used in occupational epidemiology in situations where calculation and adequate standardization of mortality rate ratios is not possible (15). The rationale for doing this is that if an occupation has a particular risk for a specific disease the proportion of deaths due to that disease will be increased relative to deaths from other causes in the group. We therefore plotted the correlation between age adjusted mortality for all causes and mortality with Covid19 across minor occupational groups for whom risk was reported in the ONS dataset, the two measures are bound to be correlated to a certain extent because all-cause mortality includes with Covid19. However, we also found a strong positive correlation between mortality with Covid19 and non-Covid19. This is consistent with findings from others which show that deaths from Covid19 closely match ‘normal’ risk within the UK population (13) and many predictors of Covid19 mortality are the same as those for others causes of death (16). Mortality from any cause amongst education professionals and teachers were towards the lower end of the distribution. For male secondary teachers, female teaching and lunchtime assistants the ratio of mortality with Covid19 to all-cause mortality was consistent with other occupational groups;female primary school teachers and female secondary school teachers appeared to be outliers in opposite directions, however the number of deaths in these groups was small and a meta-analysis across occupations working in school showed only weak evidence of heterogeneity..

### Consistency with other studies

The data we examined from ONS is consistent with the Public Health Scotland study (2) and with the Swedish study (7) which both showed similar numbers of hospitalisations due to Covid19 among teachers compared with other occupations, but our findings are also compatible with a slightly higher risk of Covid19 mortality among secondary school teachers. Mutambudzi and colleagues (17) investigated the risk of hospitalisation or death by occupation after testing positive for Covid19 among 120,075 working participants (aged 49 to 64 years) in the UK Biobank study, they found weak evidence of more hospitalisations or deaths with Covid19 among education workers compared to non-essential workers up to July 2020 (OR 1.59, 95%CI 0.87 to 2.91) after adjusting for a long list of potential confounders. However, UK Biobank is a highly selected population, which is not representative of the UK population due to a response rate of just 5% (18).

A study of data on teacher absence due to Covid19 in England during the Autumn term (September-December) 2020 found that the proportion of teachers absent due to infection appeared to be similar in both primary and secondary schools (19).The ONS schools infection survey found that primary and secondary staff had similar antibody positivity rates for SARS-CoV2 (15% and 16% respectively) at the end of Autumn term, which reflected exposure during the time when in person teaching was taking place; positivity rates among teachers were slightly lower than among all working aged adults (18%) (20). Despite this, primary school teachers in England and Wales had a lower risk of death with Covid19 compared to secondary school teachers (although confidence intervals were overlapping).

Very few studies have examined risk among teaching assistants and lunchtime assistants as separate groups; we found that despite more deaths occurring in this group in 2020 compared to the previous 5 years, they were not at increased risk of death due to Covid19 or any cause compared to working aged people. A study carried out in California used modelling to compare deaths from all causes during the pandemic and found an odds ratio for teaching assistants of 1.28, but no confidence intervals were provided for that study (21).

### Strength and limitations of the study

We used routinely collected data on mortality which includes all deaths in England and Wales, and results are therefore unlikely to be due to ascertainment bias and will be representative of the working aged population of these countries. However, we did not have access to individual level mortality data so were not able to account for potential confounders such as comorbidities or household size. For our relative risk calculations our comparison group also included school staff (although they only made up a very small proportion of the total) because we did not have age adjusted mortality rates excluding teachers. The age adjusted mortality rates calculated by ONS were based on denominators take from a survey conducted in 2019, if the school workforce has changed as a result of the pandemic these rates will be inaccurate, if any changes in the school workforce are systematically different to other professions the results will be biased. Whilst the size of the school workforce in England remained fairly stable between 2015-2019, there was a marked decrease in the number of teachers retiring (from 17,853 teachers retiring in 2015 to 12,062 teachers in 2019) (22), suggesting that the average age of teachers may have increased over this time period. We don’t know what impact the pandemic has had on the size and age distribution of those working in schools, it could be that older more vulnerable staff have retired or conversely staff may have stayed on to help with staff shortages due to teachers having to isolate. Therefore, we were unable to adequately assess the extent to which risk of death had changed during the pandemic period for these occupational groups especially among those aged over 65. In addition, the number of deaths were extremely small and subject to random fluctuations in some groups. In the period covered by this data many school staff worked remotely for large periods of time during 2020, a survey carried out by the global education company Tes found that just 22% of teachers were engaged in face-to-face teaching during the first lockdown (23),The more vulnerable staff are likely to have worked from home even when schools were fully open, which will have an impact on their workplace exposure.

## In Conclusion

Teachers, teaching and lunchtime assistants aged 20-64 years were not at high risk of death relative to the working age population in England and Wales during the Covid19 pandemic. For occupations working in schools Covid19 mortality was generally proportionate to all-cause mortality. Female primary school teachers and female secondary school teachers seemed to be outliers in opposite directions however, evidence for heterogeneity in Covid19 mortality risk among school workers was weak. There were large excesses in deaths from all causes among those working in schools aged over 65, only a third of which involved Covid19. Further research is needed to determine whether these represent numerator-denominator bias or if there is a true excesses and if so the cause of these excess deaths.

## Data Availability

The data used in the manuscript was all made publically available by the Office of National Statistics

https://www.ons.gov.uk/peoplepopulationandcommunity/healthandsocialcare/causesofdeath/bulletins/coronaviruscovid19relateddeathsbyoccupationenglandandwales/deathsregisteredbetween9marchand28december2020

## Table legends

**Table 1** Age adjusted all-cause and with Covid19 mortality per 100,000 population (95% confidence intervals) taken from 3 ONS datasets on Covid19 mortality by occupation covering different time periods during the 2020 Covid19 pandemic. Incudes the ratio of Covid19 deaths to all-cause deaths for education professionals, all professionals and all working aged people (aged 20-64 years) stratified by sex.

**Table 2** Relative risks for death occurring with Covid19 for school staff aged 20-64 years compared to the total working aged population, calculated using age-adjusted mortality rates by occupation between 9th March and 28th December 2020 from the ONS dataset.

**Table 3** Relative risks for death occurring from all causes for school staff aged 20-64 years compared to the total working aged population, calculated using age-adjusted mortality rates by occupation between 9th March and 28th December 2020 from the ONS dataset.

## Figure legends

**Figure 1** Scatterplot of mortality rate (per 100,000) for Covid19 and all causes of death for men, using age adjusted mortality rates between 9th March and 28th December 2020 from ONS.

**Figure 2** Scatterplot of mortality rate (per 100,000) for Covid19 and all causes of death for women, using age adjusted mortality rates between 9th March and 28th December 2020 from ONS

## Notes

### Competing Interest Statement

The authors have declared no competing interest.

### Funding Statement

Sarah Lewis, Caroline Relton and George Davey Smith have received a grant from National Institute for Health Research (NIHR) and UK Research and Innovation (UKRI)COVID-19 mapping and mitigation in schools (CoMMinS)

### Summary of Updates

There was a mistake in the abstract: "The mortality risk for all causes was also higher in teachers compared to all working aged people" changed to “The mortality risk for all causes was also higher in teachers compared to all professionals”. Some minor typo and wording changes were made in the discussion. A strengths and limitations section was added as per the journal requirements.

## References

1. Buonsenso, D., Roland, D., De Rose, C., Vásquez-Hoyos, P., Ramly, B., Nandipa Chakakala-Chaziya, J., Munro, A., González-Dambrauskas, S. Schools Closures during the COVID-19 Pandemic: A Catastrophic Global Situation. Preprints 2020, 2020120199 (doi: 10.20944/preprints202012.0199.v1).

2. Fenton L, Gribben C, Caldwell D, Colville S, Bishop J, Reid M, White J, Hutchinson S, Robertson C, Colhoun HM, Wood R, McKeigue PM, McAllister DA. Risk of hospitalisation with COVID-19 among teachers compared to healthcare workers and other working-age adults. A nationwide case-control study. MedRxiv 2021.02.05.21251189 [Preprint]; doi: https://doi.org/10.1101/2021.02.05.21251189

3. Helsinki Gradate School of Economics. Situation Room Report: The Corona Virus and Health Differences – In Which Socioeconomic Groups Have the Most Infections Been Observed in Finland? 19th January, 2021. https://www.helsinkigse.fi/covid19-data-en/situation-room-report-the-corona-virus-and-health-differences-in-which-socioeconomic-groups-have-the-most-infections-been-observed-in-finland/

4. Vlachos J, Hertegård E, Svalery HB. The effects of school closures on SARS-CoV-2 among parents and teachers. Proceedings of the National Academy of Sciences Mar 2021, 118 (9) e2020834118; DOI: 10.1073/pnas.2020834118

5. Ismail SA, Saliba V, Lopez Bernal J, Ramsay ME, Ladhani SN. SARS-CoV-2 infection and transmission in educational settings: a prospective, cross-sectional analysis of infection clusters and outbreaks in England. Lancet Infect Dis. 2020 Dec 8:S1473-3099(20)30882-3. doi: 10.1016/S1473-3099(20)30882-3. Epub ahead of print.

6. Falk A, Benda A, Falk P, Steffen S, Wallace Z, Høeg TB. COVID-19 Cases and Transmission in 17 K–12 Schools — Wood County, Wisconsin, August 31– November 29, 2020. MMWR Morb Mortal Wkly Rep 2021;70:136–140. DOI: http://dx.doi.org/10.15585/mmwr.mm7004e3

7. Ludvigsson JF, Engerström L, Nordenhäll C, Larsson E. Open Schools, Covid-19, and Child and Teacher Morbidity in Sweden. N Engl J Med. 2021 Jan 6:NEJMc2026670. doi: 10.1056/NEJMc2026670. Epub ahead of print. https://www.nejm.org/doi/full/10.1056/NEJMc2026670

8. Magnusson K, Nygård K, Vold L, Telle K. Occupational risk of COVID-19 in the 1st vs 2nd wave of infection. MedRxiv Preprints https://www.medrxiv.org/content/10.1101/2020.10.29.20220426v1.full.pdf

9. Coronavirus (COVID-19) related deaths by occupation, England and Wales. Release date 25th January 2021. https://www.ons.gov.uk/peoplepopulationandcommunity/healthandsocialcare/causesofdeath/datasets/coronaviruscovid19relateddeathsbyoccupationenglandandwales

10. Katikireddi SV, Leyland AH, McKee M, Ralston K, Stuckler D. Patterns of mortality by occupation in the UK, 1991-2011: a comparative analysis of linked census and mortality records. Lancet Public Health. 2017 Oct 23;2(11):e501–e512. doi: 10.1016/S2468-2667(17)30193-7.

11. Office for National Statistics. SOC2010 volume 1: structure and descriptions of unit groups. https://www.ons.gov.uk/methodology/classificationsandstandards/standardoccupationalclassificationsoc/soc2010/soc2010volume1structureanddescriptionsofunitgroups

12. Office for National Statistics. Annual population survey (APS) QMI. https://www.ons.gov.uk/employmentandlabourmarket/peopleinwork/employmentandemployeetypes/methodologies/annualpopulationsurveyapsqmi

13. Spigelhalter D. Use of “normal” risk to improve understanding of dangers of covid-19. BMJ 2020; 370: m3259. https://doi.org/10.1136/bmj.m3259

14. Office for National Statistics. Deaths registered in England and Wales. Release date July 2020. https://www.ons.gov.uk/peoplepopulationandcommunity/birthsdeathsandmarriages/deaths/datasets/deathsregisteredinenglandandwalesseriesdrreferencetables

15. Bhaskaran K, Bacon SCJ, Evans SJW, Bates CJ, Rentsch CT, MacKenna B, Tomlinson L, Walker AJ, Schultze A, Morton CE, Grint D, Mehrkar A, Eggo RM, Inglesby P, Douglas IJ, McDonald HI, Cockburn J, Williamson EJ, Evans D, Curtis HJ, Hulme WJ, Parry J, Hester F, Harper S, Spiegelhalter D, Smeeth L, Goldacre B. Factors associated with deaths due to COVID-19 versus other causes: population-based cohort analysis of UK primary care data and linked national death registrations within the OpenSAFELY platform. MedRxiv Preprints https://www.medrxiv.org/content/10.1101/2021.01.15.21249756v2.full.pdf

16. Decouflé P, Thomas TL, Pickle LW. Comparison of the proportionate mortality ratio and standardized mortality ratio risk measures. Am J Epidemiol. 1980 Mar;111(3):263–9. doi: 10.1093/oxfordjournals.aje.a112895.

17. Mutambudzi M, Niedwiedz C, Macdonald EB, et al. Occupation and risk of severe COVID-19: prospective cohort study of 120 075 UK Biobank participants. Occupational and Environmental Medicine Published Online First: 09 December 2020. doi: 10.1136/oemed-2020-106731. https://oem.bmj.com/content/early/2020/12/01/oemed-2020-106731.full#DC1

18. Tyrrell J, Zheng J, Beaumont, R. et al. Genetic predictors of participation in optional components of UK Biobank. Nat Commun 12, 886 (2021). https://doi.org/10.1038/s41467-021-21073-y

19. Southall E, Holmes A, Hill EM, Atkins BD, Leng T, Thompson RN, Dyson L, Keeling MJ, Tildesley MJ. An analysis of school absences in England during the Covid-19 pandemic. medRxiv 2021.02.10.21251484 [Preprint]; doi: https://doi.org/10.1101/2021.02.10.21251484

20. Office for National Statistics. COVID-19 Schools Infection Survey Round 2, England: December 2020. https://www.ons.gov.uk/peoplepopulationandcommunity/healthandsocialcare/conditionsanddiseases/bulletins/covid19schoolsinfectionsurveyround2england/december2020#main-points

21. Chen Y-H, Glymour M, Riley A, Balmes J, Duchowny K, Harrison R, Matthay E, Bibbins-Domingo K. Excess mortality associated with the COVID-19 pandemic among Californians 18–65 years of age, by occupational sector and occupation: March through October 2020. medRxiv 2021.01.21.21250266 [Preprint]; doi: https://doi.org/10.1101/2021.01.21.21250266

22. United Kingdom Government. Reporting Year 2019. School workforce I n England. https://explore-education-statistics.service.gov.uk/find-statistics/school-workforce-in-england#dataBlock-4189d4c5-ba35-4f48-9248-cc39f53703a2-tables

23. Tes. Reality of lockdown for school staff: 5 key findings. https://www.tes.com/news/reality-lockdown-school-staff-5-key-findings

