## Supplementary material for "Risk of death among teachers in England and Wales during the Covid19 pandemic"

**Deaths occurring among those working in schools in 2020 compared with the previous 5 years.**

**Table S1** shows a breakdown of the number of deaths by occupation in 2020 and the average for the previous 5 years among those working in schools aged 20-64 years, also includes figures for all occupations and all professionals for comparison. The number of excess deaths was calculated by subtracting the 5 year average number of deaths (provided by ONS) from the number of deaths from all causes between 9^th^ March and 28^th^ December 2020 for each occupation and occupational group. The number of excess deaths was converted into a percentage of the 5-year average (percentage excess). The percentage excess explained by Covid19 was calculated by dividing the number of Covid19 deaths by the number of excess deaths and multiplying this by 100. We set the maximum percentage excess deaths to be 100, but we did not cap the percentage excess explained by Covid19. The reason for this is to determine whether Covid19 could be a factor in some deaths which may have occurred anyway.

The number of deaths in the individual groups are mostly small and it is difficult to draw conclusions from these as they are imprecisely estimated. However, among female teachers there were fewer deaths than the 5-year average, and among male teachers deaths were 3% higher. Teaching assistants appeared to have a high number of deaths compared with the 5-year average, although only around half of these involved Covid19. When deaths across all occupations working in schools were combined, we observed 15% more deaths (excess) compared with the 2015-2019 average for the same period among men and 5% more among women. For women this is less than the excess observed across all occupations and among all professionals; for men it is similar. Deaths with Covid19 appeared to account for almost all the excess among men whereas among women there were twice as many deaths with Covid19 among school workers as there were excess deaths.

**Table S2** shows a breakdown of the number of deaths by occupation among those working in schools aged 65 and older. Excess deaths and percentage of excess deaths explained by Covid19 were calculated as for Table 1s above. This age group had much higher numbers of deaths compared with their 5-year average than the younger age group. Amongst all males in this age group there were 74% more deaths and amongst females there were 37% more than the 5-year average. However, only around a third of the excess deaths were thought to involve Covid19. The group of all professionals and of all occupations had fewer excess deaths as a proportion of their 5-year average but a higher proportion of the deaths involved Covid19 compared with those working in schools.

We did not have the denominators (total number working in each occupational group) for the figures presented in Tables 1s and 2s, so we were unable to work out the uncertainty in these estimates, and this will be very large in some of the smaller occupational groups**.**

**Comparison of risk of death with Covid19 and from all causes for education professionals versus all professionals**

**Table S3** shows relative risk of deaths occurring with Covid19 for educational professionals aged 20-64 years compared to all professionals, between 9th March and 28th December 2020. Risk of dying with Covid19 in males varied from 11.5 (95%CI 5.2 to 21.7) in 100,000 for professionals in higher education to 39.2 (95%CI 24.3 to 58.6) in 100,000 for secondary school teachers. Similarly, risk in females ranged from 9.8 (95%CI 7.5 to 12.5) in 100,000 for higher education professionals to 21.2 (95% CI 12.4 to 33.2) in 100,000 among secondary school teachers. Compared with all professionals there was strong evidence that the risk of dying with Covid19 was elevated for male (RR 2.23, 95%CI 1.54 to 3.22) and female (RR 1.66, 95%CI 1.09 to 2.51) secondary school teachers. There was also evidence that the risk of dying with Covid19 was reduced among female higher education professionals (RR 0.77, 95%CI 0.60 to 0.97). For other analyses the confidence intervals were wide, and it was not possible to draw firm conclusions, although there was a suggested reduction in risk for female primary school teachers and male further education teachers.

**Table S4** shows relative risks for all-cause mortality for educational professionals aged 20-64 years compared to the total working aged population, between 9th March and 28th December 2020. Both Male and Female higher education professionals had lower risks of mortality compared with the group of all professionals. However, there was very strong evidence that primary and secondary school teachers had higher risk of mortality from all causes compared with all professionals. This was especially true for males; male secondary school teachers had a 2.5 fold increased risk of mortality compared with all professionals and male primary school teachers a 1.8 fold increased risk.

**Ratio of mortality with Covid19 versus other causes**

We estimated the mortality due to all causes except Covid19 (Age adjusted all-cause mortality minus mortality with Covid19) and calculated the ratio of Covid19 mortality to mortality from other causes for minor occupational groups (according to ONS) and also for secondary school teachers and female primary school teachers. These figures are shown in Tables 3s and 4s.

**Table S5** shows the age adjusted mortality rates per 100,000 individuals for deaths with Covid19, plus our estimate of the non-Covid19 mortality per 100,000 among men. The ratio of deaths with Covid19 to other causes of death, ranged from 1 to 3 among male nurses and midwifes to 1 to 13 among elementary construction occupations. The ratio for male secondary school teachers was 1 death with Covid19 to 7 other deaths, which was similar to that for all education professionals.

**Table S6** shows the age adjusted mortality rates per 100,000 individuals for deaths with Covid19, plus our estimate of the non-Covid19 mortality per 100,000 among women. The ratio of deaths with Covid19 to other causes of death, ranged from 1 to 5 among welfare professionals (social workers and probation officers) to 1 to 18 among primary and nursery school teachers. The ratio for female secondary school teachers was 1 death with Covid19 to 6 other deaths. Therefore, Covid19 was less likely to be a factor in deaths among female primary and nursery school teachers than other professions and Covid19 was more likely to primary and nursery school teachers than other professions and more likely to be involved in deaths among females secondary school teachers than among other professions except probation officers and social workers. For female teaching assistants the ratio of mortality with covid to other causes was 1:10 for school midday and crossing patrol occupations this was 1:8.

Note: There were fewer occupational groups with useable data (sufficient numbers of deaths to have age-adjusted mortality risks) for women than for men, this is due to small numbers of deaths in some groups among women.

**Table S7**- We calculated the proportionate mortality rates for occupational groups working in schools for which we had sufficient numbers of Covid19 deaths (n>10). This was calculated by dividing the number of deaths involving Covid19 for the occupational group by the total number of deaths in that group. We also calculated the SE of the proportion and from that the confidence intervals for the proportion using standard formula.

We found that male secondary school teachers had a proportionate mortality rate which was very similar to all males currently in work. We found that female teaching assistants and females lunchtime assistants had proportionate mortality rates which were similar to all other females currently in work. Female secondary school teachers had a proportionate mortality rate for Covid19 which was around 50% higher than for all female workers but the confidence intervals around this estimate were wide and overlapped with the estimate for all other workers. The proportionate mortality rate for female primary school teachers was 40% lower than for all female workers and the confidence intervals for this estimate did not overlap with those for all female workers.

**Figure S1** and **S2** show scatterplots of mortality with Covid19 versus mortality from all causes for minor occupational among men (1s) and among women (2s). There was a very strong correlation between mortality with Covid19 and all-cause mortality among both men and women, correlation coefficients were 0.83 and 0.91 among men and women respectively with very small p-values. Risk of both Covid19 mortality and all-cause mortality among teachers and school staff were lower end of the scale. Male secondary school teachers fell within the centre of the plot, as did female teaching assistants and school midday and crossing patrol occupations, suggesting that the Covid19 risk reflects their normal mortality risk. However, both female primary and nursery school teachers and female secondary school teachers were slight outliers but in opposite directions, female primary and nursery school teachers had a low Covid19 risk relative to their all-cause mortality risk and female secondary school teachers had a slightly higher Covid19 risk relative to their all-cause mortality risk.

**Table S1 Deaths among working aged (20-64) people who working in education in England and Wales 9^th^ March to 28^th^ December 2020**

| Description | Men | | | | | | Women | | | | | |
| --- | --- | --- | --- | --- | --- | --- | --- | --- | --- | --- | --- | --- |
|  | Deaths involving COVID-19 | All causes of death | All causes 5 year average | Excess | % excess | Excess due to Covid19 | Deaths involving COVID-19 | All causes of death | All causes 5 year average | Excess | % excess | Excess due to Covid19 |
| Secondary education teaching professionals | 29 | 241 | 233 | 8 | 3% | 100% | 23 | 156 | 173 | -17 | -10% | NA |
| Primary and nursery education teaching professionals | 4 | 31 | 30 | 1 | 3% | 100% | 19 | 327 | 368 | -41 | -11% | NA |
| Special needs education teaching professionals | 1 | 12 | 8 | 4 | 50% | 25% | 3 | 27 | 33 | -6 | -18% | NA |
| Teaching and other educational professionals n.e.c. | 8 | 56 | 32 | 14 | 44% | 57% | 7 | 70 | 61 | 9 | 15% | 78% |
| School secretaries | 0 | 3 | 2 | 1 | 50% | 0% | 4 | 58 | 53 | 5 | 9% | 80% |
| Teaching assistants | 5 | 28 | 19 | 9 | 47% | 56% | 37 | 396 | 307 | 89 | 29% | 42% |
| Educational support assistants | 1 | 7 | 8 | -1 | -13% | NA | 3 | 55 | 58 | -3 | -5% | NA |
| School midday and crossing patrol occupations | 2 | 11 | 5 | 6 | 120% | 33% | 18 | 169 | 149 | 20 | 13% | 90% |
| All those working in schools | 50 | 389 | 337 | 52 | 15% | 96% | 114 | 1258 | 1202 | 56 | 5% | 204% |
| All professional occupations | 419 | 3144 | 2765 | 379 | 14% | 111% | 279 | 2696 | 2477 | 219 | 9% | 127% |
| All occupations | 4225 | 33904 | 29745 | 4159 | 14% | 102% | 1742 | 18419 | 16528 | 1891 | 11% | 92% |

**Table S2 Deaths among over 65s who are working in education in England and Wales 9^th^ March to 28^th^ December 2020**

| Description | Men | | | | | | Women | | | | | |
| --- | --- | --- | --- | --- | --- | --- | --- | --- | --- | --- | --- | --- |
|  | Deaths involving COVID-19 | All causes of death | All causes 5 year average | Excess | % excess | Excess due to Covid19 | Deaths involving COVID-19 | All causes of death | All causes 5 year average | Excess | % excess | Excess due to Covid19 |
| Secondary education teaching professionals | 77 | 565 | 327 | 238 | 73% | 32% | 19 | 306 | 226 | 80 | 35% | 24% |
| Primary and nursery education teaching professionals | 10 | 76 | 45 | 31 | 69% | 32% | 52 | 709 | 495 | 214 | 43% | 24% |
| Special needs education teaching professionals | 4 | 30 | 7 | 23 | 329% | 17% | 6 | 60 | 40 | 20 | 50% | 30% |
| Teaching and other educational professionals n.e.c. | 9 | 49 | 29 | 20 | 69% | 45% | 11 | 98 | 60 | 38 | 63% | 29% |
| School secretaries | 1 | 3 | 3 | 0 | 0% | NA | 13 | 99 | 88 | 11 | 13% | 118% |
| Teaching assistants | 1 | 14 | 12 | 2 | 17% | 50% | 31 | 268 | 221 | 47 | 21% | 66% |
| Educational support assistants | 1 | 6 | 5 | 1 | 20% | 20% | 13 | 64 | 48 | 16 | 33% | 81% |
| School midday and crossing patrol occupations | 4 | 13 | 6 | 7 | 117% | 57% | 46 | 325 | 231 | 94 | 41% | 49% |
| All those working in schools | 107 | 756 | 434 | 322 | 74% | 33% | 191 | 1929 | 1409 | 520 | 37% | 37% |
| All professional occupations | 637 | 4504 | 3273 | 1231 | 38% | 52% | 401 | 3480 | 2712 | 768 | 28% | 52% |
| All occupations | 7033 | 43640 | 34676 | 8964 | 26% | 78% | 2993 | 25277 | 19842 | 5435 | 27% | 55% |

**Table S3 - Covid19 related deaths for educational professionals aged 20-64 years compared to all professionals, between 9th March and 28th December 2020**

| **Description** | **Men** | | | | | **Women** | | | | |
| --- | --- | --- | --- | --- | --- | --- | --- | --- | --- | --- |
|  | Risk | Risk for all professionals | RR | 95% CI | P-value | Risk | Risk for all professionals | RR | 95% CI | P-value |
| All educational professionals | 0.00018 | 0.00018 | 1.05 | 0.82 - 1.34 | 0.74 | 0.000098 | 0.00013 | 0.77 | 0.6 - 0.97 | 0.03 |
| Higher education teaching professionals | 0.00012 | 0.00018 | 0.65 | 0.35 - 1.22 | 0.18 | NA^1^ |  |  |  |  |
| Further education teaching professionals | 0.00025 | 0.00018 | 1.40 | 0.75 - 2.62 | 0.29 | NA^1^ |  |  |  |  |
| Secondary education teaching professionals | 0.00039 | 0.00018 | 2.23 | 1.54 - 3.22 | 0.000025 | 0.00021 | 0.00013 | 1.66 | 1.09 - 2.51 | 0.017 |
| Primary and nursery teaching professionals | NA^1^ |  |  |  |  | 0.00010 | 0.00013 | 0.78 | 0.49 - 1.23 | 0.29 |
| ^1^Less than 10 deaths, rate not reported in ONS data. | | | | | | | | | | |

**Table S4 – All cause mortality for education professionals aged 20-64 years compared to all professionals, between 9th March and 28th December 2020**

| **Description** | **Men** | | | | | **Women** | | | | |
| --- | --- | --- | --- | --- | --- | --- | --- | --- | --- | --- |
|  | Risk | Risk for all professionals | RR | 95% CI | P-value | Risk | Risk for all professionals | RR | 95% CI | P-value |
| All educational professionals | 0.0015 | 0.0013 | 1.18 | 1.08 - 1.28 | 0.00023 | 0.0011 | 0.0012 | 0.92 | 0.85 - 0.98 | 0.015 |
| Higher education teaching professionals | 0.00095 | 0.0013 | 0.73 | 0.58 - 0.92 | 0.007 | 0.00073 | 0.0012 | 0.61 | 0.46 - 0.81 | 0.00059 |
| Further education teaching professionals | 0.0017 | 0.0013 | 1.29 | 1.01 - 1.65 | 0.039 | 0.00096 | 0.0012 | 0.80 | 0.62 - 1.04 | 0.092 |
| Secondary education teaching professionals | 0.0032 | 0.0013 | 2.48 | 2.18 - 2.81 | 1.41x10^-40^ | 0.0015 | 0.0012 | 1.21 | 1.03 - 1.41 | 0.018 |
| Primary and nursery teaching professionals | 0.0024 | 0.0013 | 1.84 | 1.29 - 2.61 | 0.00077 | 0.0019 | 0.0012 | 1.56 | 1.40 - 1.74 | 7.77 x 10^-15^ |

**Table S5 - Covid19 versus mortality from all other causes in men by minor occupational group**

| Group | Covid19 mortality rate | All other causes mortality rate | Ratio |
| --- | --- | --- | --- |
| Administrative Occupations: Finance | 45.9 | 226.4 | 1:4.9 |
| Administrative Occupations: Government and Related Organisations | 58.6 | 362.2 | 1:6.2 |
| Administrative Occupations: Records | 33.5 | 155.6 | 1:4.6 |
| Architects, Town Planners and Surveyors | 12.1 | 127.8 | 1:10.6 |
| Artistic, Literary and Media Occupations | 22.1 | 248.9 | 1:11.3 |
| Assemblers and Routine Operatives | 32.6 | 175.5 | 1:5.4 |
| Building Finishing Trades | 42.6 | 529.6 | 1:12.4 |
| Business, Finance and Related Associate Professionals | 15.4 | 116.5 | 1:7.6 |
| Business, Research and Administrative Professionals | 13.5 | 79.9 | 1:5.9 |
| Caring Personal Services | 91 | 379.3 | 1:4.2 |
| Construction and Building Trades | 37.3 | 392.5 | 1:10.5 |
| Construction Operatives | 31.6 | 337.4 | 1:10.7 |
| Customer Service Occupations | 42.9 | 178.3 | 1:4.2 |
| Electrical and Electronic Trades | 37.3 | 307.5 | 1:8.2 |
| Elementary Administration Occupations | 54.9 | 286.8 | 1:5.2 |
| Elementary Cleaning Occupations | 54.3 | 419.8 | 1:7.7 |
| Elementary Construction Occupations | 82.1 | 1055.7 | 1:12.9 |
| Elementary Process Plant Occupations | 143.2 | 856.1 | 1:6.0 |
| Elementary Security Occupations | 93.4 | 396 | 1:4.2 |
| Elementary Storage Occupations | 54 | 391.4 | 1:7.2 |
| Engineering Professionals | 10.8 | 84.7 | 1:7.8 |
| Functional Managers and Directors | 10.3 | 71.3 | 1:6.9 |
| Health Professionals | 23.9 | 108.9 | 1:4.6 |
| Housekeeping and Related Services | 29.1 | 220.4 | 1:7.6 |
| Information Technology and Telecommunications Professionals | 15.6 | 122.7 | 1:7.9 |
| Information Technology Technicians | 26.8 | 183.5 | 1:6.8 |
| Leisure and Travel Services | 48.4 | 257.7 | 1:5.3 |
| Managers and Directors in Retail and Wholesale | 28.3 | 133.3 | 1:4.7 |
| Managers and Directors in Transport and Logistics | 50 | 230.3 | 1:4.6 |
| Managers and Proprietors in Hospitality and Leisure Services | 72 | 421.3 | 1:5.9 |
| Managers and Proprietors in Other Services | 43.5 | 248.8 | 1:5.7 |
| Metal Machining, Fitting and Instrument Making Trades | 33.2 | 313.6 | 1:9.4 |
| Mobile Machine Drivers and Operatives | 35.1 | 363.9 | 1:10.4 |
| Nursing and Midwifery Professionals | 79.1 | 264.2 | 1:3.3 |
| Other Administrative Occupations | 25.9 | 185.8 | 1:7.2 |
| Other Drivers and Transport Operatives | 43.7 | 292.3 | 1:6.7 |
| Other Elementary Services Occupations | 59 | 360.7 | 1:6.1 |
| Other Skilled Trades | 45.8 | 329.4 | 1:7.2 |
| Plant and Machine Operatives | 82.3 | 674.8 | 1:8.2 |
| Process Operatives | 65.7 | 307.4 | 1:4.7 |
| Production Managers and Directors | 18.1 | 117.4 | 1:6.5 |
| Protective Service Occupations | 71.2 | 561.9 | 1:7.9 |
| Public Services and Other Associate Professionals | 17 | 113.3 | 1:6.7 |
| Road Transport Drivers | 57.8 | 271.2 | 1:4.7 |
| Sales Assistants and Retail Cashiers | 53.5 | 312.1 | 1:5.8 |
| Sales Related Occupations | 41.7 | 215.3 | 1:5.2 |
| Sales, Marketing and Related Associate Professionals | 22.5 | 175.5 | 1:7.8 |
| **Secondary education** | **39.2** | **283.8** | **1:7.2** |
| Teaching and Educational Professionals | 18.4 | 135 | 1:7.3 |
| Vehicle Trades | 60.2 | 441.3 | 1:7.3 |
| Welfare Professionals | 50.5 | 206.5 | 1:4.1 |

Mortality rates are per 100,000 individuals

**Table S6 -** **Covid19 versus mortality from all other causes in women by minor occupational group**

| Group | Covid19 mortality rate | All other causes mortality rate | Ratio |
| --- | --- | --- | --- |
| Administrative Occupations: Finance | 13 | 146.2 | 1:11.2 |
| Administrative Occupations: Government and Related Organisations | 19.2 | 198.8 | 1:10.4 |
| Assemblers and Routine Operatives | 39.2 | 403.8 | 1:10.3 |
| Caring Personal Services | 38.3 | 258.5 | 1:6.7 |
| Childcare and Related Personal Services | 12.8 | 120 | 1:9.4 |
| Customer Service Occupations | 13.4 | 121.6 | 1:9.1 |
| Elementary Cleaning Occupations | 20.6 | 194.5 | 1:9.4 |
| Elementary Process Plant Occupations | 49.9 | 401.6 | 1:8.0 |
| Elementary Security Occupations | 17 | 169.2 | 1:10.0 |
| Food Preparation and Hospitality Trades | 28.6 | 329.2 | 1:11.5 |
| Hairdressers and Related Services | 39.8 | 388.8 | 1:9.8 |
| Managers and Directors in Retail and Wholesale | 26.7 | 224.3 | 1:8.4 |
| Managers and Proprietors in Hospitality and Leisure Services | 35.2 | 252.8 | 1:7.2 |
| Managers and Proprietors in Other Services | 13.9 | 155.4 | 1:11.2 |
| Nursing and Midwifery Professionals | 24.7 | 162.7 | 1:6.6 |
| Other Administrative Occupations | 12 | 126.5 | 1:10.5 |
| Other Elementary Services Occupations | 21.4 | 208.5 | 1:9.7 |
| **Primary and nursery** | **10** | **177.5** | **1:17.8** |
| Sales Assistants and Retail Cashiers | 23.3 | 209.6 | 1:9.0 |
| **Secondary education** | **21.2** | **123.8** | **1:5.8** |
| **School midday and crossing patrol occupations** | **19.2** | **161.9** | **1:8.4** |
| Secretarial and Related Occupations | 10.6 | 148.1 | 1:14.0 |
| Teaching and Educational Professionals | 9.8 | 100.2 | 1:10.2 |
| **Teaching assistants** | **15** | **151.8** | **1:10.1** |
| Welfare and Housing Associate Professionals | 9.2 | 103.3 | 1:11.2 |
| Welfare Professionals | 22.6 | 120.9 | 1:5.3 |

Mortality rates are per 100,000 individuals

**Table S7- Proportionate mortality rates for those working in schools**

|  | **Occupation** | **Covid19 deaths** | **All deaths** | **Proportionate mortality rate** |
| --- | --- | --- | --- | --- |
| Men | Secondary teachers | 29 | 241 | 0.12 (0.08 to 0.16) |
|  | All other workers | 4159 | 33373 | 0.12 (0.12 to 0.13) |
| Women | Primary teachers | 19 | 327 | 0.06 (0.03 to 0.08) |
|  | Secondary teachers | 23 | 156 | 0.15 (0.10 t0 0.20) |
|  | Teaching assistants | 37 | 396 | 0.09 (0.07 to 0.12) |
|  | Midday occupations | 18 | 169 | 0.11 (0.06 to 0.15) |
|  | All other workers | 1669 | 17623 | 0.10 (0.09 to 0.10) |

**Figure S1 – Scatterplot of mortality rate (per 100,000) for Covid19 and all-cause mortality for men, between 9th March and 28th December 2020**

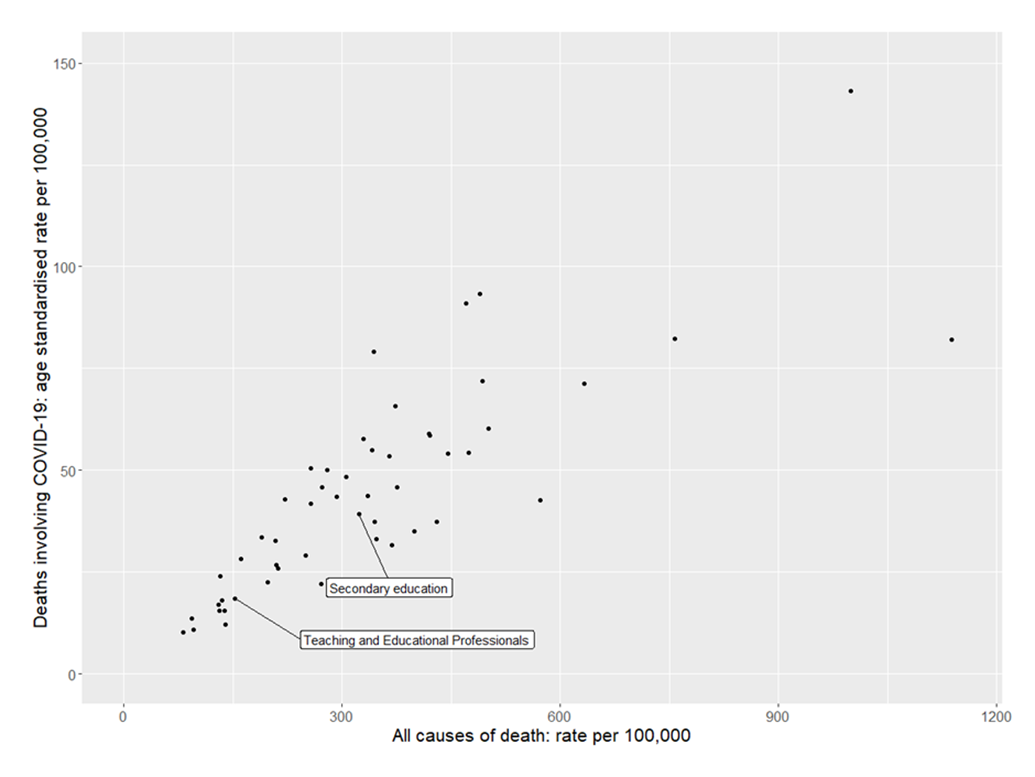

Pearson’s correlation coefficient = 0.83, *p* = 8.2 × 10^-14^

**Figure S2 – Scatterplot of mortality rate (per 100,000) for Covid19 and all-cause mortality for women, between 9th March and 28th December 2020**

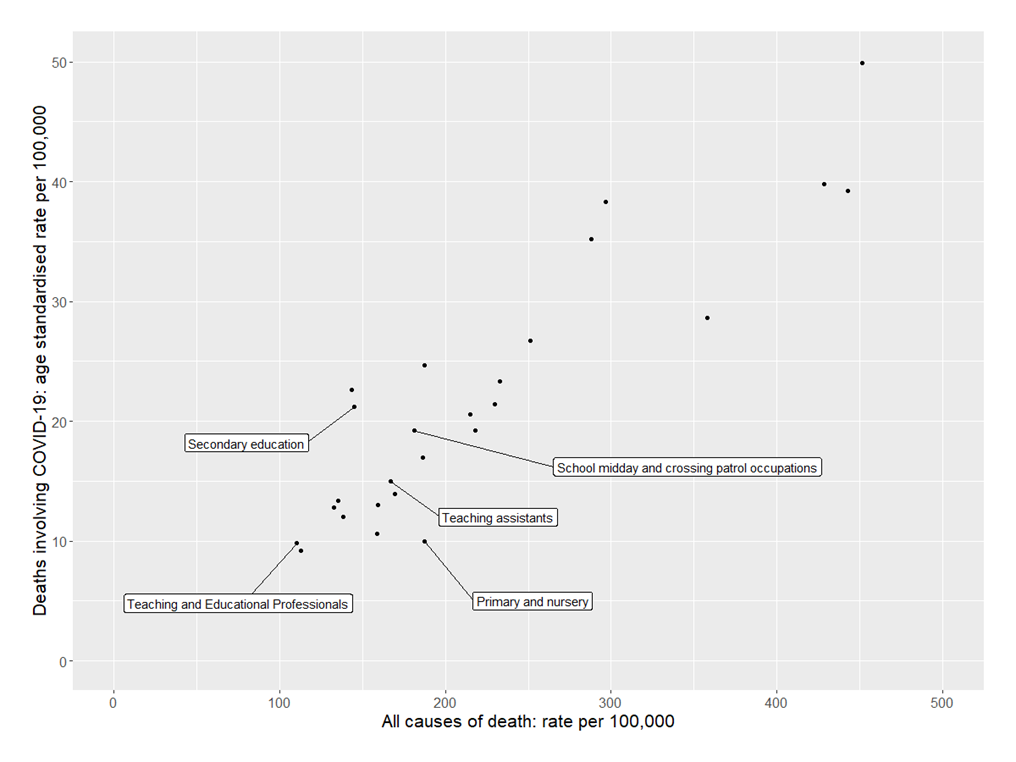

Pearson’s correlation coefficient = 0.91, *p* = 1.4. × 10^-10^
